# Optimal COVID-19 testing strategy on limited resources

**DOI:** 10.1101/2021.08.31.21262868

**Authors:** Onishi Tatsuki, Honda Naoki, Yasunobu Igarashi

**Affiliations:** Department of Pharmacoepidemiology, Graduate School of Medicine and Public Health, Kyoto University, Yoshidakonoecho, Sakyo, Kyoto, Japan; Department of Anesthesiology and Pain Clinic, Juntendo University Shizuoka Hospital, Izunokuni, Shizuoka, Japan; Department of Anaesthesiology, Tokyo Metropolitan Bokutoh Hospital, Kotobashi, Sumida, Tokyo, Japan; Laboratory for Data-driven Biology, Graduate School of Integrated Sciences for Life, Hiroshima University, Higashihiroshima, Hiroshima, Japan; Theoretical Biology Research Group, Exploratory Research Center on Life and Living Systems (ExCELLS), National Institutes of Natural Sciences, Okazaki, Aichi, Japan; Laboratory of Theoretical Biology, Graduate School of Biostudies, Kyoto University, Yoshidakonoecho, Sakyo, Kyoto, Japan; E2D3.org, izumi-cho, Kokubunji, Tokyo, Japan; Center for Research on Assistive Technology for Building a New Community, Nagoya Institute of Technology, Nagoya, Aichi, Japan

## Abstract

The last three years have been spent combating COVID-19, and governments have been seeking optimal solutions to minimize the negative impacts on societies. Although two types of testing have been performed for this—follow-up testing for those who had close contact with infected individuals and mass-testing of those with symptoms—the allocation of resources has been controversial. Mathematical models such as the susceptible, infectious, exposed, recovered, and dead (SEIRD) model have been developed to predict the spread of infection. However, these models do not consider the effects of testing characteristics and resource limitations. To determine the optimal testing strategy, we developed a testing-SEIRD model that depends on testing characteristics and limited resources. In this model, people who test positive are admitted to the hospital based on capacity and medical resources. Using this model, we examined the infection spread depending on the ratio of follow-up and mass-testing. The simulations demonstrated that the infection dynamics exhibit an all-or-none response as infection expands or extinguishes. Optimal and worst follow-up and mass-testing combinations were determined depending on the total resources and cost ratio of the two types of testing. Furthermore, we demonstrated that the cumulative deaths varied significantly by hundreds to thousands of times depending on the testing strategy, which is encouraging for policymakers. Therefore, our model might provide guidelines for testing strategies in the cases of recently emerging infectious diseases.

## 1 Introduction

The Coronavirus disease 2019 (COVID-19) emerged in Wuhan, China, raising concerns regarding global healthcare [1,2]. By April 2020, the COVID-19 Alpha variant pandemic had infected 5.5 million people, and 350,000 people had died, owing to its high aerosol transmission ability and the lack of specific treatment in the early stages [3]. Medical resources in hospitals were primarily used to treat COVID-19 patients [1,2]. As of April 2020, approximately 10% of hospital beds, or 10–20% of ICU beds were occupied with COVID-19 care [3–5]. Moreover, in May 2020, the COVID-19 Beta variant emerged. The society needed to be updated about the variant of concern (VOC) such as the Beta, Gamma, Delta, and Omicron variants every time a new variant emerged [6–8].

To minimize the number of deaths, society must be aware of the advantages and disadvantages of COVID-19 testing [9]. From an individual perspective, testing has advantages in that asymptomatic infected individuals can be detected and prepared for symptomatic treatment, whereas from a societal perspective, testing prevents secondary infections, expecting a reduction in the number of deaths [10–12]. The Alpha variant pandemic in April 2020, in which no specific treatment was established and testing characteristics, that is, sensitivity and specificity, were unknown, illustrates the drawbacks of testing, particularly in the early pandemic stage. From an individual perspective, the testing result had no impact on medical care because there was no specific treatment, but from a societal perspective, testing was performed aimlessly, and people were uncertain about the testing outcomes, resulting in the wastage of medical and human resources. Therefore, policymakers must consider the testing characteristics when determining the volume of testing at each early stage of an emerging VOC.

The testing policies to minimize the number of deaths in the early stages of the COVID-19 Alpha variant pandemic were controversial [7,10,11,13,15–20], and the controversy was centered on the two extreme policies for balancing the medical supply and demand: mass-testing and no-testing [21]. According to the mass-testing policy, everyone must be tested for public health, regardless of their symptoms [22–24]. The mass-testing policy assumes that testing and hospitalization of asymptomatic patients are important for reducing the overall death rate even in the absence of a specific treatment. Conversely, the no-testing policy claims that testing must be limited to symptomatic patients [21]. According to the no-testing policy, asymptomatic patients cannot expect to benefit from mass-testing in the absence of a specific treatment. Despite differences in these two policies’ assumptions, they both support testing on people with symptoms. However, these approaches disagree regarding the size of the tested asymptomatic population.

What testing strategy is most practical for minimizing the number of deaths? There are two testing strategies for people without symptoms: follow-up-testing-dominant strategy, which follows up and tests the exposed population, and mass-dominant testing strategy, which randomly tests the infected population. Uncertainty about how follow-up and mass-testing of asymptomatic populations will affect the number of deaths and determine the worst and optimal outcomes, particularly in the early stages of emerging VOC in the future, remains a challenge [7,10,11,13,15–20].

In this study, we developed a testing-SEIRD model, aiming to evaluate a testing strategy that combines follow-up and mass-testing in terms of minimizing the number of deaths during the early stages of the emerging VOC. The testing-SEIRD model considers the testing characteristics, testing strategies, hospitalized subpopulation, and the amount of medical resource [25]. Using this model, we examined the optimal and worst testing strategies under the assumption that medical resources are both infinite and finite. We found that the optimal testing strategy significantly depends on the cost ratio between mass and follow-up testing. Therefore, this study provides insights into how to minimize the number of deaths in the absence of a specific treatment during the early stages of a pandemic.

## 2 Model

To examine the impact of testing on the infection population dynamics, we developed a novel model by incorporating a hospitalized subpopulation, testing strategy, and testing characteristics into the classical SEIRD model. Generally, the subpopulation susceptible dynamics, exposed, infectious, recovered, and dead people best summarize the SEIRD model (Fig. 1A) as follows:

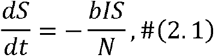

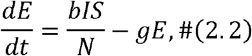

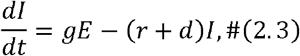

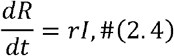

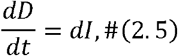

where *S, E, I, R*, and *D* indicate the populations of susceptible, exposed, infectious, recovered, and dead people, respectively; *N* indicates the total population, that is, *N*=*S*+*E*+*I*+*R*; *b* indicates the exposure rate, which reflects the level of social activity; and *g, r*, and *d* indicate the transition rates among the subpopulations. In this model, the recovered population is assumed to acquire permanent immunity, indicating that they will never be infected.

**Figure 1:**
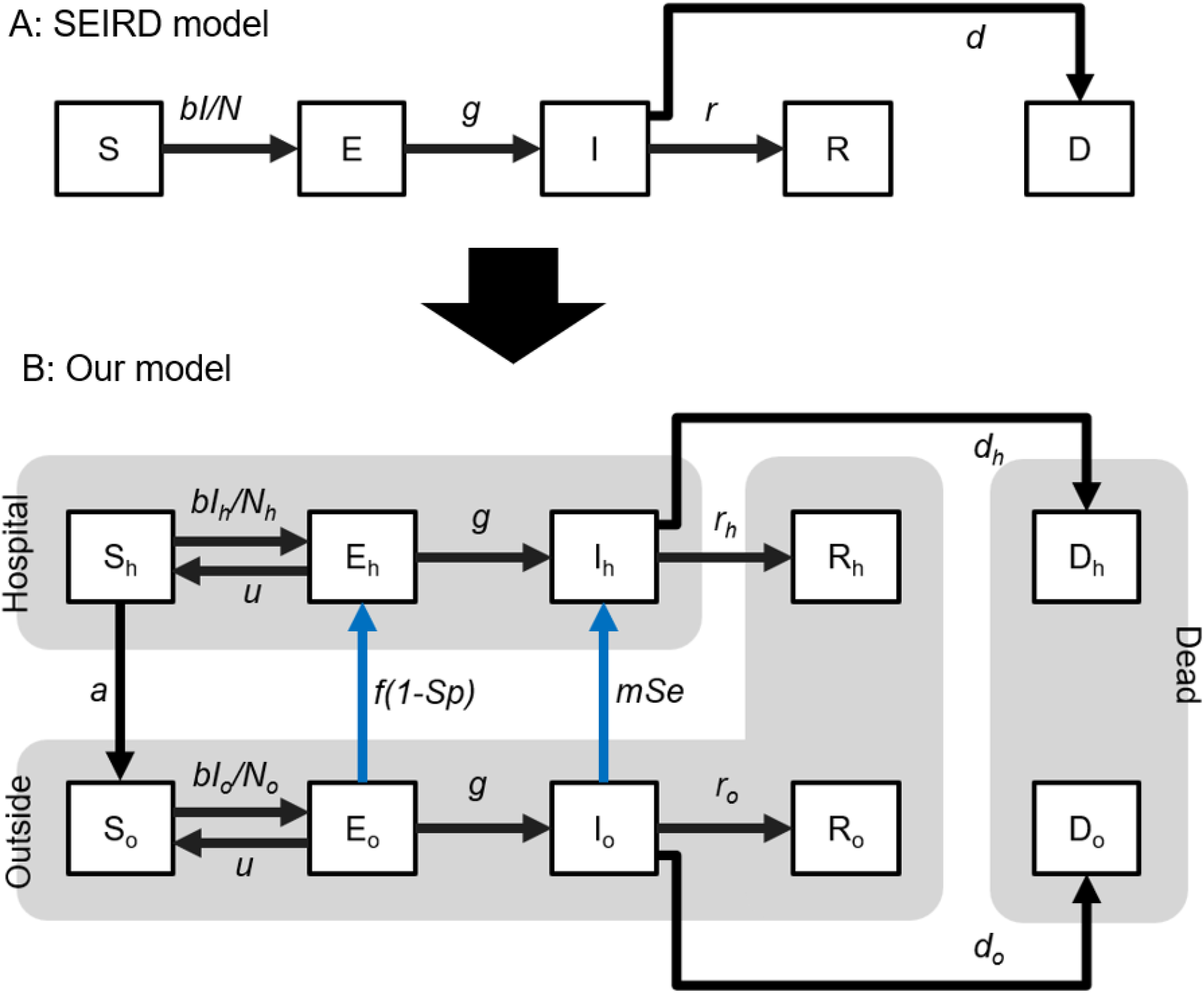
Schematic of the classical SEIRD and testing-SEIRD models. (A) Classical SEIRD model: An infectious population “I” exposes a susceptible population “S” at a rate inversely proportional to the infectious population. The exposed population “E” becomes infectious “I.” The infected population finally recovers “R” or is dead “D.” (B) Testing-SEIRD model: The population is divided into two subpopulations; inside and outside the hospital. The exposed “E_o_” and the infectious population outside “I_o_” are hospitalized if evaluated as positive after testing. A susceptible population “S_h_” remains at the hospitals. The black lines indicate population transitions, regardless of the capacity effect. The blue lines indicate population transition, considering the capacity effect. Transitions from “E_o_” to “E_h_” and “I_o_” to “I_h_” are categorized as hospitalized.

To incorporate the testing characteristics and testing strategies into the classical SEIRD model, we divided the population into outside and inside of the hospital (Fig. 1B). The dynamics of the population outside the hospitals are described using the following:

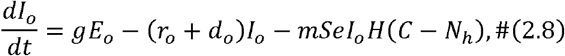

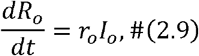

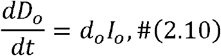

and those inside hospitals are described using the following:

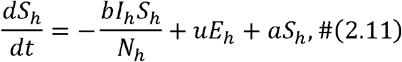

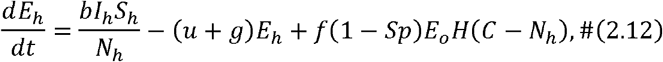

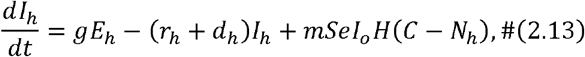

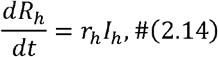

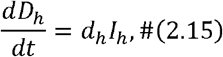

where *X*_*o*_ and *X*_*h*_ indicate each population outside and inside the hospital (*X* ∈{*S, E, I, R, D, N*}), respectively; *N*_*o*_ and *N*_*h*_ indicate the total populations outside and inside hospitals, respectively (that is, *N*_*o*_=*S*_*o*_+*E*_*o*_+*I*_*o*_+*R*_*o*_+*R*_*h*_ and *N*_*h*_=*S*_*h*_+*E*_*h*_+*I*_*h*_); *a* indicates the rate of discharge of *S*_*h*_ from the hospital to the outside; *u* and *g* indicate the non-infection and infection rates, respectively; *C* indicates the capacity of hospitals. We assumed that the nature of the disease would determine these parameters; making them independent of hospitals both inside and outside. *r*_*j*_ and *d*_*j*_ (*j*∈{*o, h*}) indicate the recovery and death rates from infection, respectively, where *r*_*o*_ < *r*_*h*_, and *d*_*h*_ < *d*_*o*_; *f* and *m* indicate the rates of follow-up and mass-testing, corresponding to the extent to which health centers follow exposed populations and take-up infected populations having symptoms, respectively; *Sp* and *Se* indicate specificity and sensitivity, respectively, as testing characteristics. The model assumed that *I* has a fixed proportion of symptomatic and asymptomatic individuals, and that symptomatic infected individuals receive mass-testing. The sigmoid function *H*(*x*) = 1/(1+exp(*x*)) introduced the hospitalization capacity. The parameter values and initial conditions are listed in Table 1 and discussed in the Materials and Methods section.

**Table 1:**
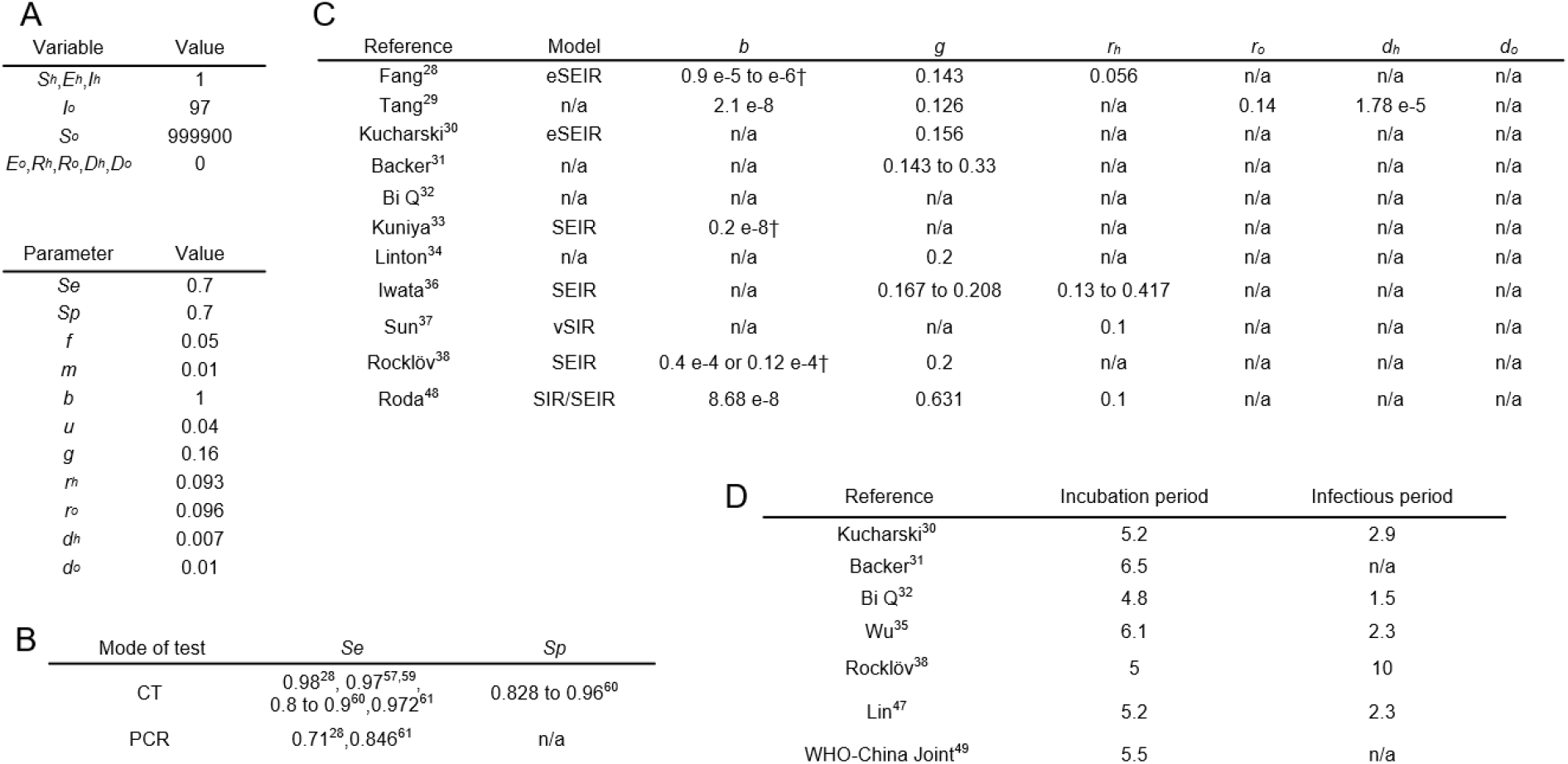
Variables and parameters in reports during the early stages of the pandemic. (A) Initial values for variables and parameters, (B) Reported sensitivity and specificity of the polymerase chain reaction (PCR) and CT for detecting COVID-19, Cells expressed as n/a indicate that we could not find the (C) reported transition parameters using models. Values with † are calculated from the original values for comparison. All values have a [one/day] dimension. We could not find values or models for the cells expressed as n/a. The values with † equal original values are divided by the total population, and (D) Reported incubation period and infectious periods. Each value has a [day] dimension.

## 3 Results

First, we examined the basic behavior of the testing-SEIRD model using simulations, as shown in Fig. 2. Similar to the classical SEIRD model, the infection primarily expands, and infectious populations (I_h_ and I_o_) transiently increase in response to the presence of infectious people. Susceptible populations (S_h_ and S_o_) gradually decrease and change into recovered populations (R_h_ and R_o_) through the exposed (E_h_ and E_o_) and infectious (I_h_ and I_o_) states. During this process, the number of dead people increases gradually, as shown in Fig. 2A. Because of hospital overcrowding, the outside and hospitalized populations decrease and increase in response to testing, respectively, and their time courses are affected (Fig. 2B). The outside and hospitalized populations are divided into five types of populations (susceptible, exposed, infectious, recovered, and dead) (Figs. 2C and 2D). According to Fig. 2E, daily reports of positive tests and deaths transiently increase with different peak timings, and the peak of positive tests precedes that of deaths.

**Figure 2:**
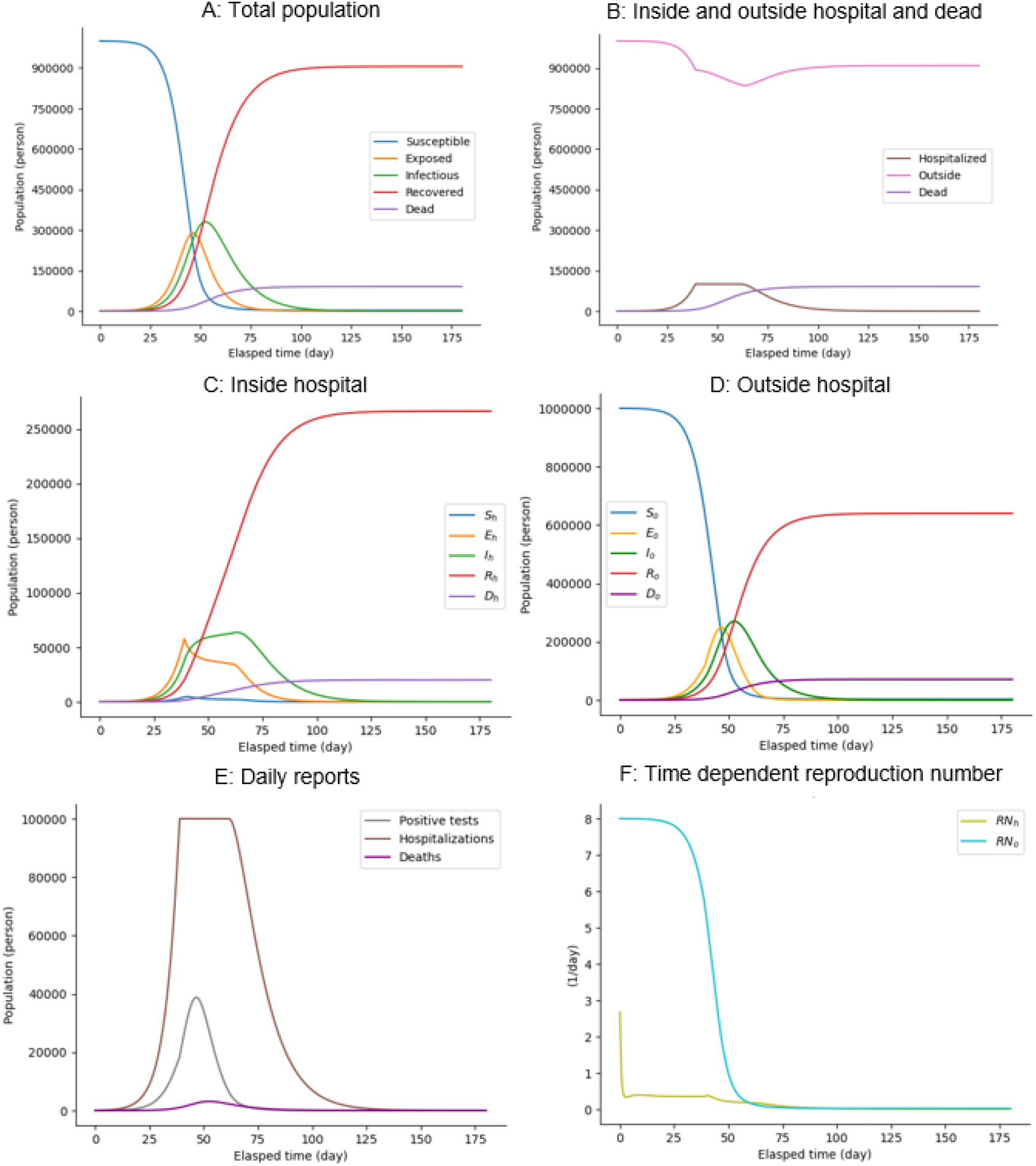
Changes in components over time in the testing-SEIRD model. Time-courses of (A) populations of all infectious states, irrespective of being inside and outside hospitals; (B) populations inside and outside hospitals and dead populations, irrespective of infectious states; (C) populations of all infectious states inside hospitals; (D) populations of all infectious states outside hospitals; (E) Daily reports of positive test results, hospitalizations, and deaths; and (F) Time-courses of reproduction numbers inside and outside hospitals, as described in Materials and Methods section.

To evaluate the speed of an infectious outbreak, we computed the basic reproduction number *RN*, which is the expected number of infections caused by one infected person until recovery (see Materials and Methods). Reproduction numbers outside hospitals, *RN*_*o*,_ switches from greater than one to less than one around the peak timing of infectious populations outside (Fig. 2F). Conversely, reproduction numbers inside hospitals, *RN*_*h*,_ are less than one around the peak timing. This indicates that the infectious population in hospitals increases owing to outside factors rather than an infectious spread within the hospitals. The testing-SEIRD model recapitulates the basic infection dynamics of the total population as observed in the classical SEIRD model (Fig. 2A) and enables us to examine the effect of the testing strategy and testing characteristics with different populations inside and outside hospitals.

To investigate the impact of hospitalization capacity on infection dynamics, such as daily reports of positive test results, hospitalizations, and deaths, we simulated the testing-SEIRD model with various capacities (Figs. 3A–3C). The results demonstrate that as the capacity increases, the maximum positive tests, maximum hospitalizations, and cumulative deaths linearly decrease, increase, and decrease, respectively. They all plateau at approximately 30% capacity (Figs. 3D–3F), and notches are observed to reflect the capacity effect (Figs. 3B, 3D, 3E, 3F, 3G, 3H, and 3I). Additionally, we examined their peak timings and found that they changed nonlinearly within certain time window ranges (Figs. 3G–3I). These results suggest that the capacity change has a significant effect on the disease’s rate of spread but only a minor effect on timing.

**Figure 3:**
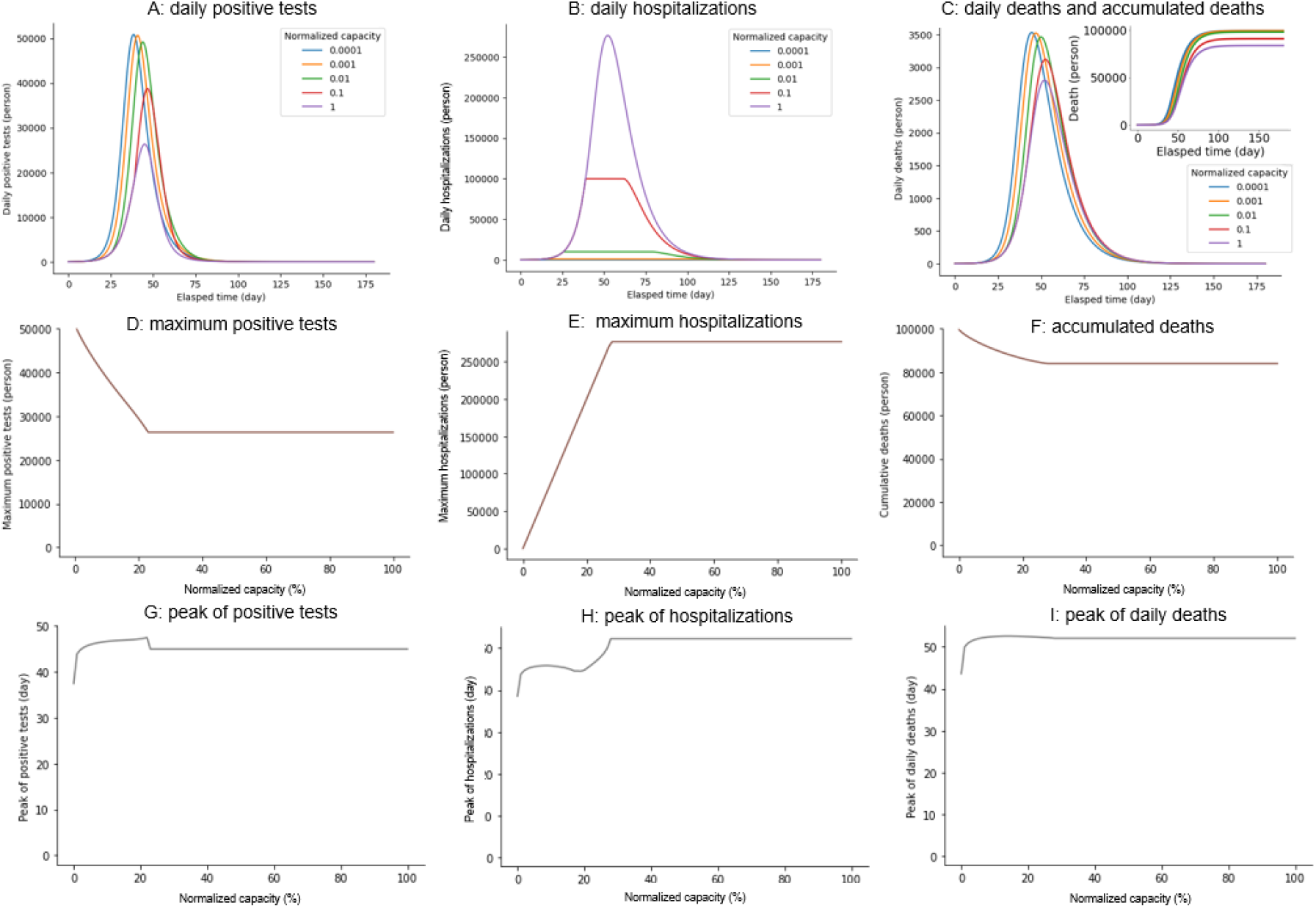
Impact of hospitalization capacity on the three variables. Time courses of (A) Daily reports of positive test results. (B) Daily reports of hospitalizations. (C) Daily reports of deaths with varying hospitalization capacity. C/N indicates the capacity normalized to the total population. Hospitalization capacity dependencies of (D) Maximum-positive reports. (E) Maximum hospitalizations. (F) Cumulative deaths. Hospitalization capacity-dependencies of (G) Peak of daily reports of positive tests. (H) Peak of hospitalizations. (I) Peak of daily deaths.

To illustrate the impact of the testing strategy on infectious outcomes, we examined the cumulative deaths, maximum number of positive tests and hospitalizations, varying follow-up, and mass-testing rates. The infectious spread shows an all-or-none response depending on the testing strategy (red and blue regions in Fig. 4). Sensitivity analyses demonstrated the robust maintenance of such a profile regardless of the model parameters (Figs. S1 and S2). The number of cumulative deaths was almost constant with a small amount of both the follow-up and mass-testing (red region in panels in the first row of Fig. 4A); however, the combination of follow-up and mass-testing successfully suppressed the infectious disease spread (blue region in the panels in the first row of Fig. 4A). Furthermore, the maximum number of hospitalizations was immediately saturated by either the follow-up or mass-testing because of the limited hospitalization capacity (panels in the first row in Fig. 4B). The maximum number of positive tests increased more quickly with follow-up testing compared with mass-testing (panels in the first row in Fig. 4C). According to statistics, the number of cumulative deaths varied significantly depending on the strategies; there was a 724-fold difference between the 90596 and 125 deaths at the optimal and worst strategies with a 1:1 cost ratio for follow-up to mass-testing. Other infectious outcomes also depend on the strategies: there was a 466-fold difference between 49424 and 106 hospitalizations and a 250-fold difference between 96525 and 135 daily positive tests with the same cost ratio.

**Figure 4:**
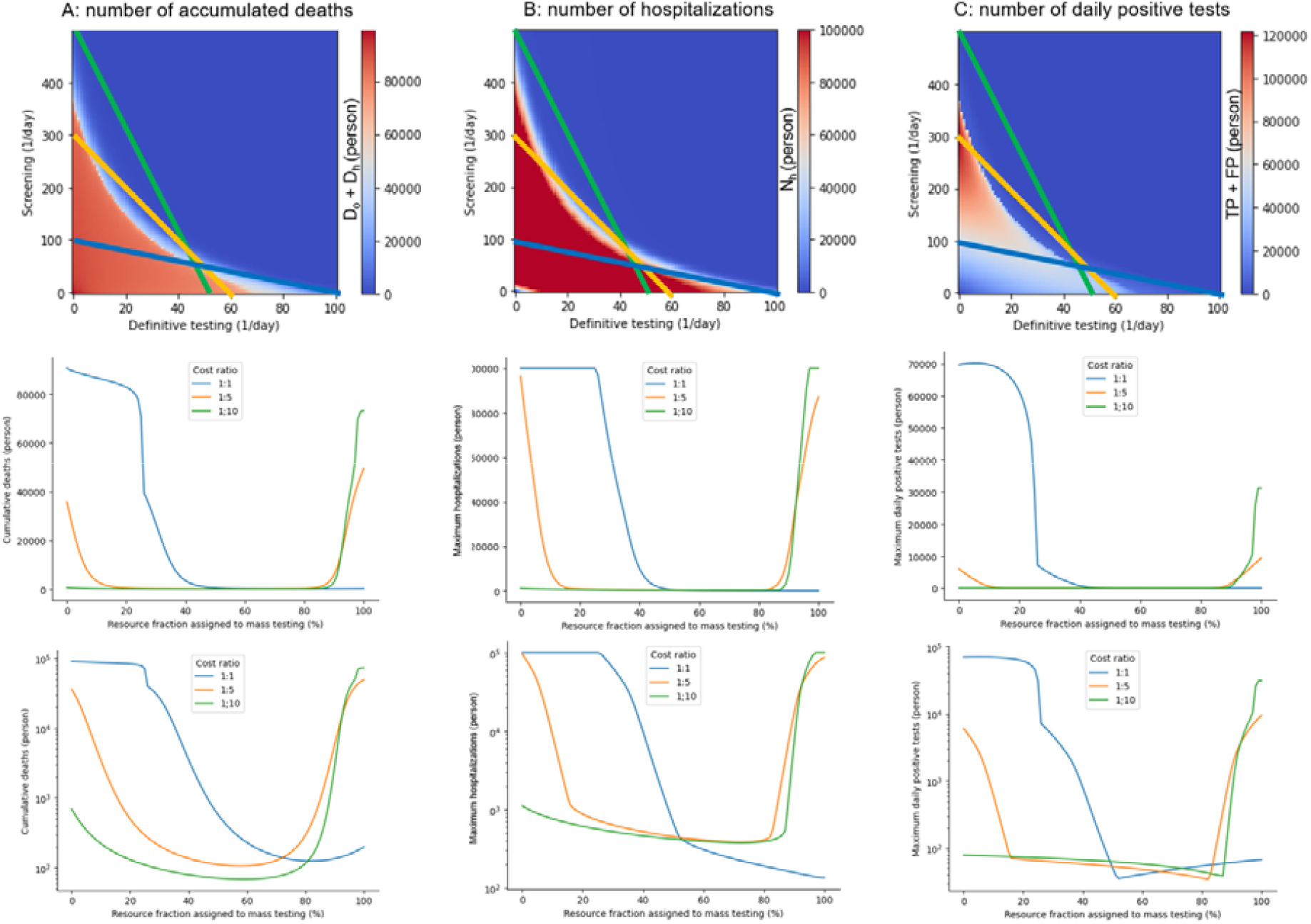
Infectious spread based on the testing strategy. The panels in the first row represent the number of (A) cumulative deaths, (B) maximum hospitalizations, and (C) maximum daily positive tests depending on the rates of follow-up and mass-testing. The three lines in these heatmaps represent the possible testing strategies subject to different total resources for testing with different ratios for the testing costs. *L*=500, *c*_*f*_ =1, and *c*_*m*_ =10 in the green line; *L*=300, *c*_*f*_ =1, and *c*_*m*_ =5 in the blue line; and *L*=100, *c*_*f*_ =1, and *c*_*m*_ =1 in the orange line. The panels in the second row represent the numbers along the three lines in the heatmaps. The panels in the third row represent semilog-plots of the second row.

Subsequently, realistic scenarios were considered adapting to the limited resource *L*. Practically, the follow-up and mass-testing rates cannot be controlled because of the limited medical resources for both follow-up and mass-testing. Therefore, it is necessary to determine the amount of resources allocated to the follow-up and mass-testing. Here, we consider all the possible decisions subject to the limited resource *L* as follows:

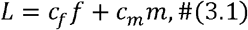

where *c*_*f*_ and *c*_*m*_ indicate the costs for follow-up and mass-testing, respectively; *f* and *m* indicate the extent of follow-up and mass-testing. We illustrated three lines using various *L, c*_*f*_, and *c*_*m*_, based on the disease, economic, and technological situations of each country (panels in the first row of Fig. 4). The three colored lines in the heat maps correspond to settings that are *L*=500, *c*_*f*_ =1, and *c*_*m*_ =10 in the green line; *L*=300, *c*_*f*_ =1, and *c*_*m*_ =5 in the blue line; and *L*=100, *c*_*f*_ =1, and *c*_*m*_ =1 in the orange line, respectively. Given the total amount of resources, we selected the optimal testing strategy on the line represented by Equation (3.1). We demonstrated that the worst decisions (that is, the choice of *f* and *m*) significantly varied depending on the situation (panels in the last row of Fig. 4).

Regarding the high resources and low ratio of the cost of follow-up testing to that of the mass-testing cost, the number of cumulative deaths abruptly increases as the resource fraction of mass-testing exceeds 90% (green line in Fig. 4A). This indicates that the mass-dominant testing is the worst strategy for minimizing the cumulative deaths. Conversely, the number of cumulative deaths abruptly decreases at the resource fraction of 20–30% (blue line in Fig. 4A) assigned to mass-testing owing to low resource availability and a high ratio of follow-up to mass-testing costs. Contrary to the previous case, this result suggests that follow-up-dominant testing is the worst strategy. Regarding the intermediate situation between the two cases above, the simulation showed a U-shape with the resource fraction assigned to mass-testing ranging from approximately 10–80% (orange line in Fig. 4A). These results suggest that both follow-up and mass-dominant testing strategies should be avoided. The choice of f and m also changed in the profiles of maximum hospitalizations and positive reports (Figs. 4B and 4C). The optimal strategy for each country/region depends on resource availability.

Moreover, we examined the effects of the testing characteristics (that is, sensitivity and specificity) on the three variables (that is, the number of cumulative deaths, hospitalizations, and positive tests). We conducted sensitivity analyses for *Se* and *Sp* using values ranging from zero to four in 0.01 increments. We obtained almost the same heatmaps in the sensitivity-specificity space although the heatmaps were inverted along the x-axis (Fig. 5). The Equations (2.7), (2.8), (2.12), and (2.13) reveal that sensitivity and one-specificity essentially play the same roles in the follow-up and mass-testing. The sensitivity and specificity of the test cannot be changed, whereas the testing strategy can be arbitrary. If the sensitivity is low, an increase in the mass-testing rate can produce the same infectious result with high sensitivity. Conversely, if the specificity is low, a decrease in the follow-up testing rate can produce the same infectious result with high specificity. Therefore, we must manage the optimal testing strategy based on the testing sensitivity and specificity that cannot be changed.

**Figure 5:**
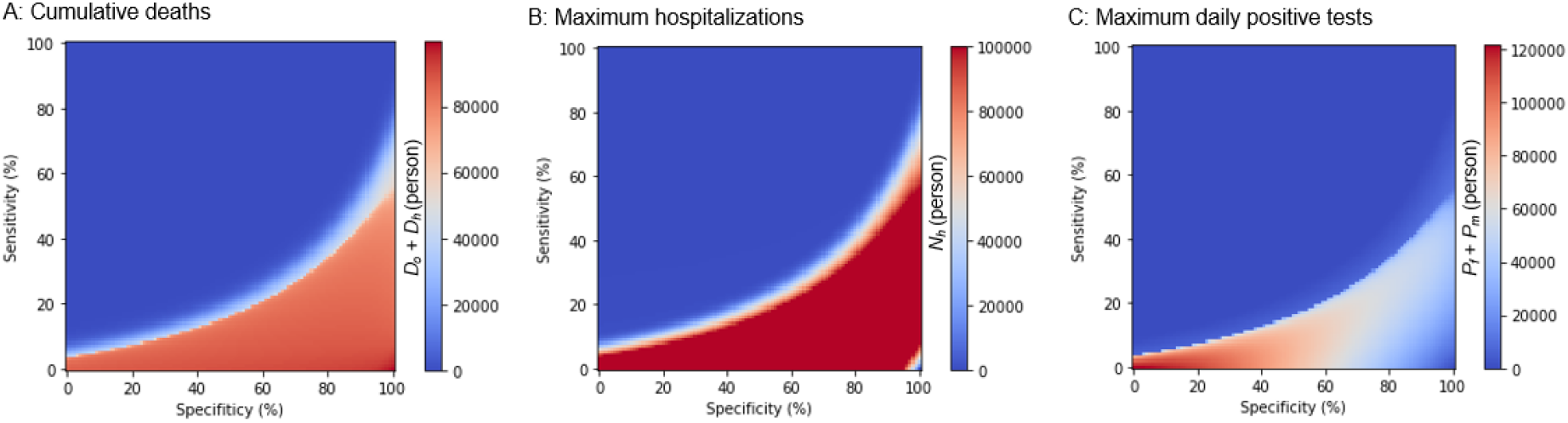
Infectious spread based on the testing properties. Numbers of (A) Cumulative deaths, (B) Maximum hospitalizations, and (C) Maximum daily positive tests based on the sensitivity and specificity of the testing.

We investigated how the infection is spread based on the testing strategy. However, this is from the viewpoint of a perfect observer who knows the exact timeline of the latent populations. Practically, we were unable to determine all the model variables, such as the exposed and infectious populations inside and outside hospitals; however, we could merely monitor positive reports by follow-up and mass-testing. In this study, we verified whether these two types of positive reports reflect the latent infectious population, which is the most resource-consuming and challenging social issue. Using regression analysis (see Materials and Methods), we demonstrate that latent infectious populations can be predicted from daily positive reports of follow-up and mass-testing (Figs. 6A–6C). These results suggest that the infectious population is not only proportional to the total number of follow-up and mass-testing positive results but also proportional to their weighted sum (Fig. 6D). There are some situations where weights can be negative, depending on the model parameters. We found that follow-up testing’s weight for positive reports was negative with high positive predictive values. This is because the negative weight of *P*_*f*_ represses the estimates of the latent number of infectious people, reflecting a low positive predictive value (Fig. 6D).

**Figure 6:**
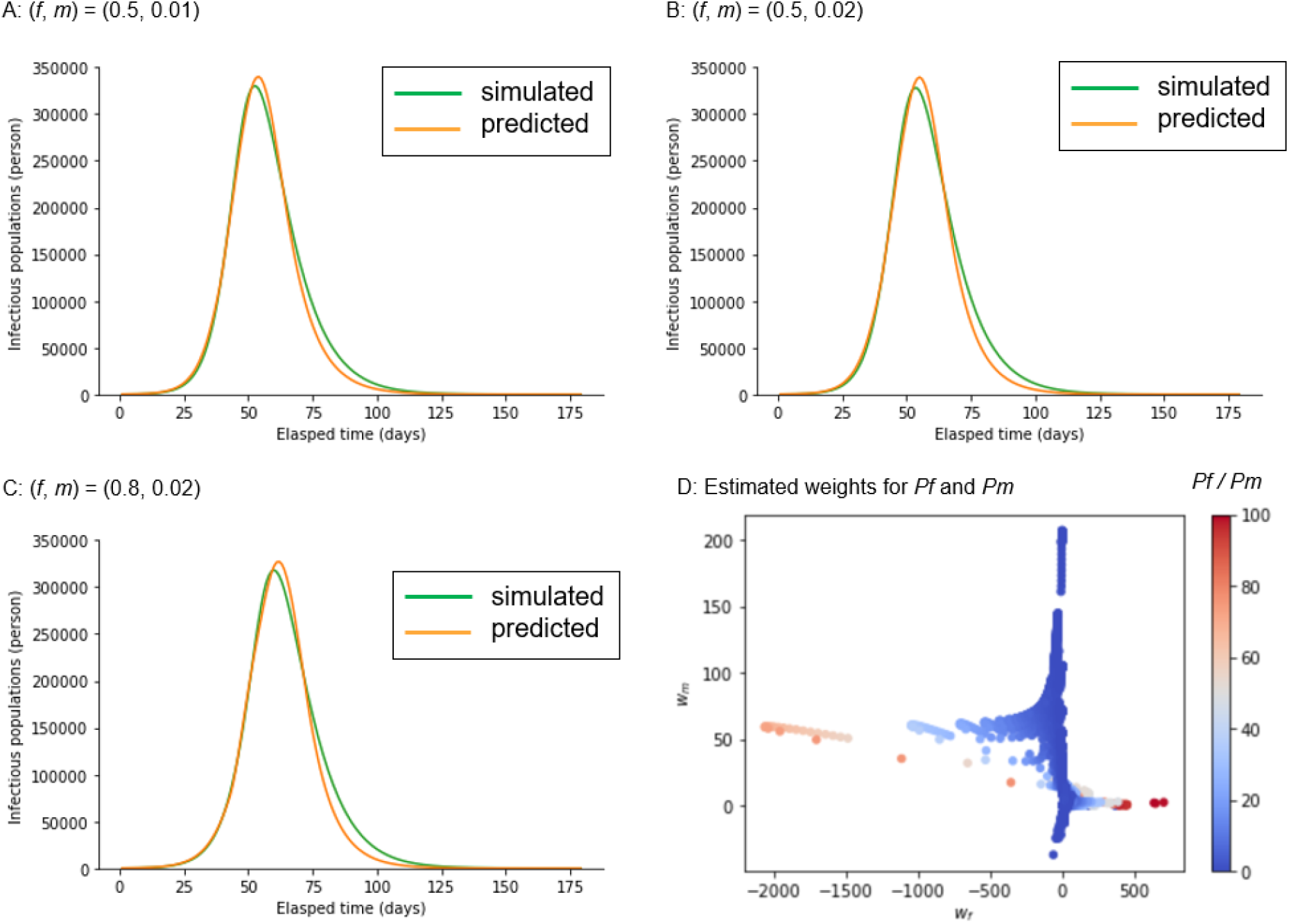
Prediction of infectious population from daily reports of positive test results. (A-C) The Green and orange lines indicate the simulated and predicted infectious populations (I_h_ and I_o_) with different testing strategies. The linear regression *as w*_*f*_*P*_*f*_ + *w*_*m*_*P*_*m*_, where *w*_*f*_ and *w*_*m*_ indicate the weights and *P*_*f*_ and *P*_*m*_ indicate the daily positive reports of follow-up and mass-testing, namely, *f* (*1-Sp*) and *mSe*, respectively, was used to estimate the infectious populations. The least square method was used to estimate the weights. (D) The estimated weights for *P*_*f*_ and *P*_*m*_ are plotted, considering various combinations of ratios of the follow-up cost to the mass-testing cost (*P*_*f*_ /*P*_*m*_).

## 4 Discussion

### Conclusion

We developed a testing-SEIRD model with two discrete populations inside and outside hospitals, the impact of testing strategy (follow-up testing [*f*], and mass-testing [*m*]), and testing characteristics (sensitivity [*Se*] and specificity [*Sp*]) on three important variables (the number of maximum positive tests, maximum hospitalizations, and cumulative deaths (Fig. 1)). By simulating the model with parameters representing the early stages of the COVID-19 Alpha variant pandemic, we demonstrated that the optimal and the worst testing strategies are subject to limited medical resources (Fig. 4). Additionally, we highlighted the possibility that the infectious population can be predicted by a weighted sum of positive follow-up and mass-testing reports (Fig. 6).

#### 4-1 Related work

Infectious dynamics models, such as SEIRD models and their alternatives, which have been widely used for policy making through model simulation, are abundant [1,26-46]. Although some of the previous models included a hospital compartment [1,26-28,30,34], they did not consider the testing strategy and testing characteristics. Our model assumes that in certain models, the exposed people do not infect the susceptible ones but they end up being affected [1,26-38]. All models, except the model with intervention strategies, [39] did not consider the testing cost. Similar to our model, three studies modeled the control of infectious outbreaks, which addressed the possibility of an optimal solution for controlling infectious outbreaks [39], the stable situation depending on the proportion of the susceptible population [40], and the basic reproduction number depending on contact rate [43]. However, to the best of our knowledge, no model has been developed that considers the effects of both testing characteristics and limited medical resources on the number of deaths. Consequently, our testing-SEIRD model introduced new factors: the hospital compartment, testing strategy, testing characteristics, and medical resources, compared with the previous SEIRD model (Figs. 2–4). The testing-SEIRD model also comprehensively encompasses the classical SEIRD model, which corresponds to the condition where f and m are both zero.

#### 4-2 Model prediction

Our model has three advantages. First, the testing-SEIRD model provides the optimal testing strategy for various situations. The model provides heatmaps based on the three variables’ numbers in the space of the testing strategy (Fig. 4). These heatmaps indicate the best direction, which is shown by the blue region in Fig. 4. This corresponds to the settling of infections using the shortest path. Second, the testing-SEIRD model can predict the optimal and worst strategies, considering the limited medical resources and ratios for the testing costs (Fig. 4). Because the total costs of medical resources and testing depend on the country, our model provides an optimal testing strategy unique to each country. Third, the testing-SEIRD model demonstrates that the latent number of infectious populations can be predicted from daily positive reports of the follow-up and mass-testing (Fig. 6).

#### 4-3 Validity of the model components

Here, we discuss the validity of the model components, which is not factored by the previous models. First, we focus on the transition from E_o_ to E_h_ (Fig. 1). We assume that the follow-up testing causes the hospitalization of the exposed population. Populations who have only recently been exposed but have not yet developed symptoms do not participate in the tests. They only test when the follow-up encourages them. Second, in relation to the transition from I_o_ to I_h_, we assume that the mass-testing causes the hospitalizations of the infectious population, which is defined as a person with symptoms. In our model, we address the rate of mass-testing as a modifiable parameter because the rate depends on the volume of tests, such as PCR and the degree of social penalty if it is positive. Third, we consider the transition from E_o_ to S_o_ and E_h_ to S_h_. In our model, all the exposed populations are not necessarily infected and some return susceptible compared with the previous models, which assume that all exposed populations are destined to be infected [28,30,32,33,35-38,47-49]. Consistent with our model, some exposed populations return to susceptible populations without developing symptoms. Finally, because the above-mentioned assumptions regarding exposure, infection, and hospitalization processes are common in VOCs, our model is not specific to the Alpha variant but is applicable to other VOCs [8]. Combining new components and the testing-SEIRD model is consistent with the previous simulation model and reflects and incorporates a practical viewpoint.

#### 4-4 Validity of the model parameters

We used parameters from earlier reports before the Beta variant emerged in South Africa in May 2020 [8] (Table 1) because the earlier reports contained homogeneous Alpha variant data. After May 2020, the reports present an inhomogeneous mixture of multiple variants. A sensitivity analysis was performed after setting the sensitivity and specificity of testing to 0.7 each, as shown in (Fig. 4). The results were robustly guaranteed. The incubation and infectious periods remained roughly stable in VOCs, while the number of reproductions and mortality rates differed among variants [8,12,14]. A sensitivity analysis of *b* and *u* provided a robust guarantee for the number of reproductions and reinfections [42] effects (Fig. S1). Mortality was considered in the model with *d*_*o*_ and *d*_*h*_ and these values were sensitivity analyzed (Fig. S2). A sensitivity analysis robustly guaranteed *a* or the rate of discharge from S_h_ (Fig. S3). Although these values are based on the COVID-19 Alpha variant, our sensitivity analysis indicates that the testing-SEIRD model robustly generated the optimal and worst testing strategies for other VOCs with different parameters.

Our model does not assign a specific value to the basic reproduction number even though it is one of the most crucial variables in infectious diseases [39,51,52]. Instead, it is only obtained using Equations (5.1) to (5.3). This is permissive because the reproduction number depends on the exposure rate (*b*) [43], and we performed a sensitivity analysis for the value of *b* (Fig. S1).

#### 4-5 Future studies

Considering the future perspectives of our model, first, our testing-SEIRD model only simulates an infection’s single peak time course. However, we observed several COVID-19 infection peaks in many countries [53]. To incorporate the multiple peaked dynamics, we must introduce the socio-psychological effects caused by policies such as lockdown and social distancing. Second, our model assumes that all populations are homogeneous and does not address stratification based on attributes such as gender, age, social activities, and comorbidities [54,55]. Future research should consider this perspective. Finally, our model did not include the effects of vaccination. There are current efforts to fight the spread of COVID-19 using messenger RNA (mRNA) vaccines. Our results appear favorable; however, we do not know the duration of the effect of the vaccinations or the effectiveness of the acquired immunity against VOCs [53,56,57]. Therefore, the tug-of-war between the evolution of vaccines and the spread of virus remains elusive.

## 5 Materials and Methods

### 5-1 Parameter set

The parameters and initial conditions of the simulation are listed in Table 1A. We used parameters from the COVID-19 Alpha variant studies. The total population *N* was set to 1,000,000 according to the United Nations statistical papers: The World’s Cities in 2018 states that one in five people worldwide live in a city with more than one million inhabitants, and the median value of inhabitants is between 500,000 and one million [58]. Therefore, sensitivity *Se* and specificity *Sp* were both set to 0.7, corresponding to those of the PCR for detecting COVID-19 (Table 1B) [28,47,58-61]. The values of *b, g, r*_*h*_, *r*_*o*_, and *d*_*h*_ are based on previous reports (Table 1C) [3,29–34,36–38,50]. The sum of *u* and *g* is the inverse of the incubation period during the exposed state, which is reportedly five days (Table 1C) [31–33,49,60]. The sum of *r* and *d* is the inverse of the infectious period during the infectious state, which is reportedly ten days (Table 1D) [31,32,35,60].

### 5-2 Definitions of reproduction numbers

We computed the time courses of the reproduction numbers inside and outside hospitals (*RN*_*h*_ and *RN*_*o*_) using Fig. 2.

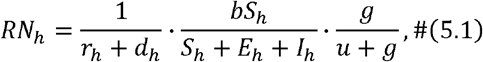

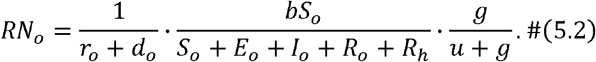

Here, the first, second, and third factors in these equations indicate the average infectious period, infection rate, and probability that the exposed state transits to the infectious state, respectively. The reproduction number in the classical SEIRD model was defined in previous studies [1,27–34] as follows:

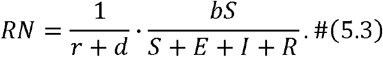

### 5-3 Code and data availability

All codes and data required to reproduce the results of this study are hosted in Github at https://github.com/bougtoir/testing-SEIRD. The Github repository contains Jupyter notebooks for Runge-Kutta method differential equations and their visualization. The python codes described in the Jupyter notebooks can reproduce all figures in this study without the need for external files or settings.

## Data Availability

All relevant data are within the study and its supporting information files.

https://github.com/bougtoir/testing-SEIRD

## Acknowledgments

We thank Tomohiko Takada M.D. (Ph.D.) and Yoshika Onishi M.D. (Ph.D.) for providing the basic concepts of clinical NNT. We thank Yoshiaki Yamagishi M.D. (Ph.D.), Tomokazu Doi M.D. (Ph.D.), and Tatsuyoshi Ikenoue M.D. (Ph.D.) for revising the early manuscript. We also acknowledge Prof. Hiroshi Nishiura for organizing a summer boot camp in 2014 to provide fundamental knowledge on infectious disease modeling.

## Ethics

This study did not involve human or animal subjects.

## Author Contributions

O.T. and Y.I. conceived the initial ideas. O.T. developed and implemented the method, processed, and analyzed the data, and wrote the initial draft of the manuscript. H.N. revised the initial draft of the manuscript and reviewed the method. Y.I. supervised the project. All authors contributed to the final writing of the manuscript.

## Supporting information

**Figure S1:**
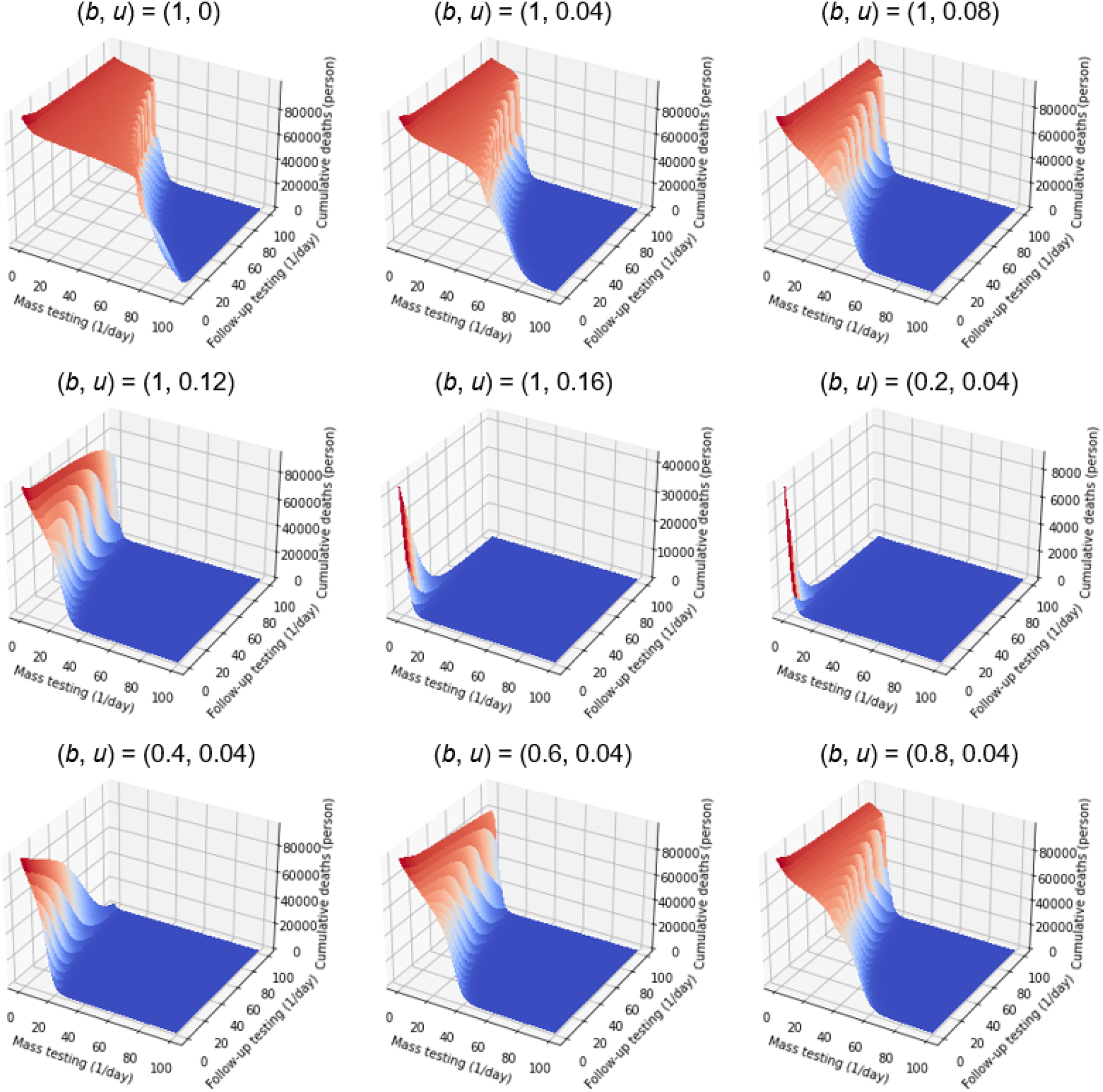
Sensitivity analyses of parameters *b* and *u* on the number of cumulative deaths. Simulations were performed using different values of *b* and *u*.

**Figure S2:**
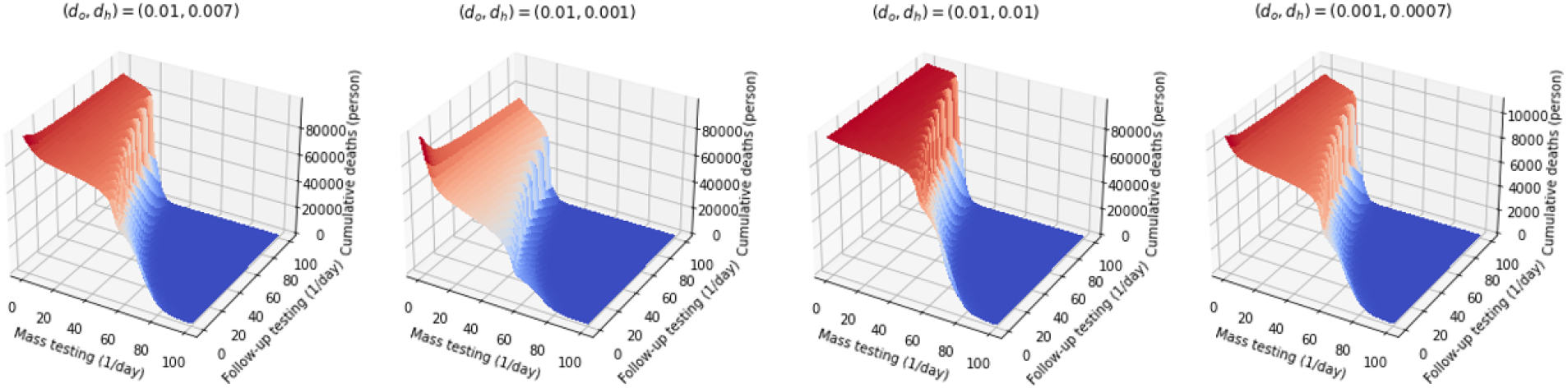
Sensitivity analyses of *d*_*o*_ and *d*_*h*_ on the number of cumulative deaths. Simulations were performed using different values of *d*_*o*_ and *d*_*h*_, where (*d*_*o*_, *d*_*h*_) of (0.01, 0.007) is a reference standard; (0.01, 0.001) means advance in treatment; (0.01, 0.01) means futile treatment; and (0.001, 0.0007) means reduction in overall mortality.

**Figure S3:**
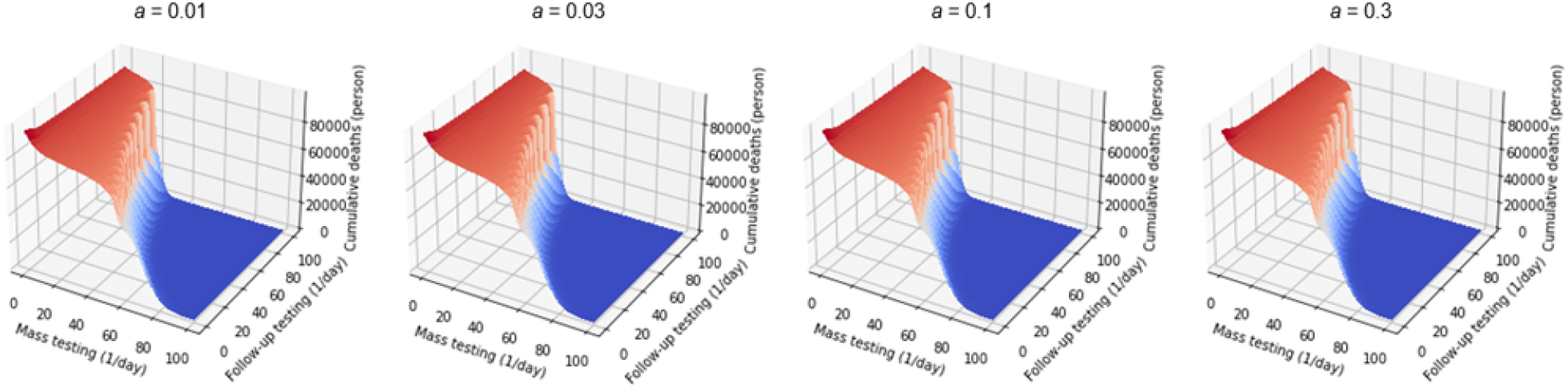
Sensitivity analyses of parameter *a* on the number of cumulative deaths. Simulations were performed using different values of parameter *a*.

